# Discovery and Verification of Extracellular microRNA Biomarkers for Diagnostic and Prognostic Assessment of Preeclampsia at Triage

**DOI:** 10.1101/2023.01.26.23285072

**Authors:** Robert Morey, Lara Poling, Srimeenakshi Srinivasan, Carolina Martinez-King, Adanna Anyikam, Kathy Zhang-Rutledge, To Cuong, Abbas Hakim, Marina Mochizuki, Kajal Verma, Antoinette Mason, Vy Tran, Morgan Meads, Leah Lamale-Smith, Mariko Horii, Gladys A. Ramos, Peter DeHoff, Mana M. Parast, Priya Pantham, Louise C. Laurent

**Author notes:** xThese authors contributed equally to this manuscript (Robert Morey, Lara Poling).

## Abstract

Preeclampsia (PE) is a major cause of maternal and neonatal morbidity and mortality. Current methods for evaluation of patients with suspected PE do not reliably predict which patients will later develop PE nor distinguish between PE with and without severe features. The objective of this study was to identify candidate extracellular miRNA (ex-miRNA) biomarkers for early diagnosis and prognosis of PE in a cohort of subjects presenting for evaluation of suspected PE. Small RNA-seq libraries were created from maternal serum samples, prospectively collected from participants undergoing evaluation for suspected PE between 20-40 weeks gestational age. Measurements for bivariate biomarkers, consisting of ratios of pairs of ex-miRNAs were performed. 110 bivariate ex-miRNA biomarkers passed both discovery (48 cases, 34 controls) and verification (23 cases, 18 controls) criteria. Specific biomarkers differed in their patterns of expression among different categories of hypertensive disorders in pregnancy (HDP). An iterative machine learning method was then used to identify 3 bivariate miRNA biomarkers that, when applied serially, differentiated cases from controls with a sensitivity of 93% and a positive predictive value (PPV) of 55%, and additionally distinguished between PE cases of different severity. In an independent validation cohort of 11 cases and 7 controls, these three biomarkers had a sensitivity of 91% and a PPV of 85%. We have discovered, verified, and validated 3 bivariate ex-miRNA biomarkers, which, when applied serially, enable accurate early diagnosis of preeclampsia. Bivariate ex-miRNA biomarkers can distinguish between different subtypes of HDP.

## Introduction

Preeclampsia (PE) is a form of placental dysfunction that affects approximately 5-8% of pregnancies worldwide(1, 2). PE manifests as hypertension and proteinuria, and in severe cases can lead to end-organ injury(1, 2). Currently, the only intervention that can halt the progression of disease is delivery, and thus many pregnancies affected by preeclampsia are delivered early to protect the health of the mother and fetus. As a result, preeclampsia is the leading cause of iatrogenic preterm birth.

Although it is thought that PE arises from abnormal trophoblast differentiation and invasion in early placental development(3–5), clinical manifestations do not arise until the second half of pregnancy. The most common symptom of PE is headache, followed by visual changes and abdominal pain. Typically, the diagnosis of PE is made when a pregnant patient displays new-onset hypertension (blood pressure > 140/90 mm Hg) and proteinuria (> 0.30 g/24 h)(6), but atypical cases where one of these features is absent or uninformative are not uncommon, particularly for women with preexisting conditions, such as chronic hypertension or kidney disease. A subset of patients with PE develop additional clinical features, including liver or kidney dysfunction, low platelets, cerebral edema, seizures, or placental abruption. Given the broad array of clinical presentations for PE, it can be difficult to differentiate the early signs and symptoms of PE from other conditions.

Currently, early diagnosis of and/or risk-assessment for the later development of PE is problematic due to the lack of assays that are highly specific for this disease. Accurate evaluations are important when planning the intensity of pregnancy surveillance or determining the timing of delivery. If delivery is induced too early, the child may be unnecessarily exposed to complications associated with prematurity. However, if the decision to deliver is made too late, the mother and child may be exposed to an increased risk of severe manifestations of PE, which can lead to serious morbidity or death. This dilemma frequently leads to expensive and lengthy hospital stays for patients at high risk of PE. The management of PE could be substantially improved, and unnecessary hospitalizations could be avoided, if an assay were developed to rapidly and accurately diagnose early PE and/or assess the risk of severe PE.

At present, a highly sensitive and specific assay for early diagnosis or prognostic evaluation of PE does not exist(7), although multiple commercial protein biomarker tests for PE have been reported. The most accurate of these tests are designed to detect two proteins in the blood; placental growth factor (PlGF) and soluble FMS-like tyrosine kinase 1 (sFlt-1). PlGF is important in the development of a healthy placenta, as it promotes the formation of new blood vessels, while sFlt-1 inhibits the function of PlGF(8). In women with PE, PlGF levels may be decreased, while sFlt-1 levels may be increased(8). The level of these two proteins is reported as a ratio of sFlt-1:PlGF. As the ratio of sFlt-1:PlGF increases, so does the risk of preterm PE.

The primary limitation of the sFlt-1:PlGF ratio is its relatively low positive predictive value, which numerous studies have reported to be between 30-40%(9– 11). Still, it is currently the most promising protein biomarker screening tool for PE. The benefit of the sFlt-1:PlGF ratio is its negative predictive value, which is helpful in ruling out PE in and identifying women at low-risk for development of PE among those with signs or symptoms of PE. While the sFlt-1:PlGF may be helpful in certain scenarios, the diagnosis of PE still depends on standard clinical signs and symptoms. Studies have yet to demonstrate that pregnancy outcomes are improved by screening suspected cases of PE using the sFlt-1:PlGF ratio(8).

To date, the FDA has not approved a test for PE based on the measurement of sFlt-1:PlGF ratio. Additionally, these tests have been excluded from the PE screening practices recommended by the United States Preventative Services Task Force, the National Institute for Health and Care Excellence, the American College of Obstetricians and Gynecologists, and the Society of Obstetricians and Gynecologists of Canada. Collectively, these organizations find limited clinical utility in the commercially available PE biomarker tests and have requested the development of better screening biomarkers and tools for risk prediction(7).

To improve upon existing assays, many research efforts have evaluated proteins other than sFlt-1 and PlGF(12– 20). Some studies have moved beyond protein biomarkers altogether to nucleic acid-based laboratory tests. These studies initially focused on long non-coding RNA molecules, cell free DNA, and cellular mRNA(21–24), but over the past decade, an increasing number of studies have explored the utility of extracellular microRNAs (ex-miRNAs) as diagnostic or prognostic biomarkers for PE(25–42).

Many of these biomarker studies have utilized a candidate approach, measuring levels of specific biomolecules based on results of prior studies demonstrating that they are expressed specifically by the placenta and/or play a role in placenta development or function. While this approach has yielded significant results in several published studies, none of these studies have led to the development of a clinical assay. Thus, the use of nucleic acid biomarkers for PE is still in the early phase, and their roles in the pathophysiology of PE are still not understood.

Since it has not been established that only placental biomolecules are perturbed in women at elevated risk of developing PE, we reasoned that a comprehensive unbiased approach might reveal novel biomarkers for PE. Therefore, the primary aim of this study was to use an unbiased transcriptomic approach to discover ex-miRNA biomarkers for the early diagnosis of PE in a cohort of pregnant women being evaluated for signs and symptoms of PE. Post-hoc analysis was also performed to explore the use of ex-miRNA biomarkers for prediction of severity of PE in those who developed this complication.

## Methods

### Study design

Patients were consented under an IRB protocol approved by the Human Research Protections Program at UC San Diego. Serum samples and the RNA samples isolated from them were labeled with study identifiers with no personal identifiable information. Each subject signed a HIPAA release and agreed to have demographic and clinical data collected from the electronic medical record (EPIC) throughout the course of pregnancy and postpartum. The primary objective of the study was to use an unbiased transcriptomic approach to discover ex-miRNA biomarkers for the early diagnosis of PE in a cohort of patients at high risk of developing PE. A secondary objective was to find ex-miRNA biomarkers that predict the severity of PE in those who developed this complication. Our sample size was determined by the number of patients consented during a given time frame and data inclusion and exclusion criteria are detailed below and in the text. Patients were randomly assigned to the Discovery and Verification groups and investigators performing the laboratory analysis were blinded to patient’s diagnosis.

### Recruitment and enrollment

Patients being evaluated in the UC San Diego OB Triage unit, between 20^0^-40^6^ weeks (20 weeks 0 days-40 weeks 6 days) gestational age, presenting with suspected PE (e.g., complaint of headache, visual disturbances, epigastric pain, hypertension, proteinuria, or fetal growth restriction) were approached and screened for enrollment. Patients who received a diagnosis of PE prior to delivery were classified as cases, while all patients who did not develop PE were classified as controls. All diagnoses were determined via retrospective review of the medical charts at least 8 weeks post-delivery.

#### Inclusion Criteria

- Pregnant women age *≥* 18
- Prenatal care at UC San Diego Prenatal Care Clinics
- Singleton pregnancy
- Gestational age between 20^0^-40^6^ weeks gestational age
- Undergoing evaluation for suspected PE

#### Exclusion Criteria

- Previous admission during the current pregnancy for suspected PE
- Previously diagnosed during the current pregnancy with severe preterm PE
- Plan for immediate delivery
- Admission to the hospital for other indications, with post-admission development of signs or symptoms requiring evaluation for PE

### Adjudications of Pregnancy Outcomes

The clinical outcome of each pregnancy was adjudicated by two Obstetrician-Gynecologists, at least one of which was board-certified in Maternal Fetal Medicine.

### Sample Collection

After informed consent was obtained, 10 mL blood was collected by peripheral venipuncture, allowed to clot in an upright position for at least 20 minutes at room temperature, and then centrifuged at 2000xg for 10 minutes. The serum was removed and placed into aliquot tubes each containing 0.5 mL serum and stored at 80°C. Sample processing was completed within 2 hours of blood collection. Extracellular RNA (exRNA) isolation and analysis was performed retrospectively, post-delivery. Thus, exRNA levels were not known by the subject, physician, or study team during the pregnancy and had no impact upon the evaluation and management of the subject. 131 subjects were recruited and donated samples for the initial discovery and verification groups. An additional 18 subjects were recruited and donated samples to verify candidate bivariate biomarkers.

### Laboratory Analysis

exRNA was isolated from each serum sample using the Plasma/Serum Circulating and Exosomal RNA Purification Kit (Slurry Format) (Norgen Biotek Corp, Ontario, Canada). Quality control of isolated RNA was performed using the Agilent RNA 6000 Pico Kit (Agilent, Santa Clara, California). Small RNA-seq libraries were constructed using the NEBNext® Multiplex Small RNA Library Prep Set for Illumina® (New England Biolabs, Inc., Ipswich, MA). The libraries were then cleaned using the DNA Clean & Concentrator(tm)-5 Kit (Zymo Research, Irvine, California) and quality control of the libraries was performed using the Agilent High Sensitivity DNA Kit (Agilent, Santa Clara, California). Equal volumes of the libraries were pooled, size-selected for products that were 120-135 nucleotides in length using a Pippin Prep with a 3% agarose gel cassette (Sage Science, Beverly, Massachusetts), and run on a MiSeq instrument at the UC San Diego Institute for Genomic Medicine (IGM) Genomics Core using the MiSeq Nano Reagent Kit (Illumina, San Diego, California). Samples that produced adequate numbers of miRNA read counts were then rebalanced to produce similar numbers of miRNA reads and sequenced on a HiSeq 4000 instrument to produce 1×75 bp reads (Illumina, San Diego, California) at the UC San Diego IGM Genomics Core.

### Sequencing Data Analysis

Small RNA-seq data were trimmed and mapped to known human sequences using the exceRpt Small RNA-seq Pipeline Workflow implemented in the Genboree Workbench(43). Each sample was evaluated for miRNA read depth and miRNA complexity. Samples that exhibited low miRNA read depth (<500,000 miRNA reads) or low miRNA complexity (<300 different miRNA species) were excluded from analysis. miRNAs were filtered such that miRNAs with at least ten raw reads in at least 50% of cases or controls were retained, resulting in 267 pass-filter miRNAs. The filtered and scaled sequencing data were normalized by global scaling and visualized using the Qlucore Omics Explorer Software (Qlucore, Lund, Sweden) and evaluated using the statical analysis methods described below. miRNA tissue contributions were determined using the recently reported fractional contribution of each cell/tissue type to each miRNA(44). For each miRNA, the tissue with the maximum contribution and any tissue within 10% of the maximum was considered to be a contributing tissue source.

### PEER

Small RNA-seq data was filtered such that miRNAs were required to have reads detected in at least 25% of the patients. This resulted in an input dataset of 619 miRNAs. Cases and controls were divided among Discovery (training) and Verification groups aiming to maintain a similar gestational age range in both groups. The raw counts for the remaining miRNAs are provided in (**Supplementary Data file 7**). Starting with the discovery group, read counts were log2 transformed and loess normalized. The PEER package (v. 1.0)(45) was then run to model the effect of our cases and controls. The PEER process performance was assessed using principal component analysis and Kruskal-Wallis test. Using the PEER processed data, a p-value comparing the cases and controls was calculated for each miRNA using a generalized linear model and Chi-square test. AUCs were then generated for each miRNA with the pROC package, using the Delong and bootstrap methods to establish confidence intervals and a t-test to generate p-values(46). As previously published(44), bivariate datasets were created with all possible miRNA ratios and ranked by an inverse rank sum using 1000 bootstraps. At each iteration, AUCs were calculated in a similar manner to the univariate data, as well as the squared correlation between the miRNA ratio and the binary case/control column, and the mean difference between cases and controls. Five ranks were derived from the resulting 1000 iterations for each miRNA ratio: 1) the mean of the AUCs; 2) the lower 25% quantile of the AUCs; 3) the mean of the squared correlation; 4) the upper 75% quantile of the AUCs; and 5) the p-value calculated using a t-test. Each rank was then inverted and summed for each miRNA ratio to obtain a final ranking (**Supplementary Data file 8**). The same procedure was used for processing the verification group. Analysis was performed using R 3.4.1. Clustering of bivariate biomarkers was done using the scipy hierarchical, agglomerative clustering package using a weighted method and a custom Pearson correlation distance metric. The median, z-scaled bivariate ratios were used as input into the clustering algorithm. Enrichment analysis of the 110 bivariate biomarkers was done using the cumulative distribution function (CDF) of the hypergeometric distribution using the top 10,000 bivariate biomarkers as the initial population size.

The Discovery and Verification datasets were processed separately. Ranked values were combined with the calculated statistics for each of the miRNA ratios. The data was then filtered for only miRNA ratios with an AUC > 0.5 and then sorted by their calculated rank. The top 1000 ranked miRNA ratios were then assessed in the verification dataset. We considered any miRNA ratio that had a lower 25% quantile AUC over 0.7 in the verification dataset “verified” (110 miRNA ratios). To assess the performance of our method, we performed principal component analysis on both the set of verified miRNA ratios and 110 randomly selected ratios. The ratio values used in the principal component analysis were obtained by combining the discovery and verification datasets before performing normalization. Combined discovery and verification miRNA ratio ranking values, the raw AUCs, the squared correlations, and pvalues the ranks were based on, the mean and median of both the cases and controls for the separate miRNAs in the ratio, and the normalized ratio values from the combined dataset for the top 1000 miRNA ratios in the discovery dataset are provided in (**Supplementary Data file 9**).

The univariate discovery and verification dataset were created separately using the AUC’s and Chi-square pvalues generated for each of the 619 miRNAs and then ranked using the Chi-square values. The top 100 ranked miRNAs from the univariate discovery dataset were extracted from the univariate verification dataset and ranked based on the Chi-square p-value. To assess the performance of our method, we performed principal component analysis using all patient samples on all 619 miRNA’s, the top 100 discovery miRNAs, and the top 10 miRNAs based on the verification dataset ranking. Normalized univariate data combined with calculated AUCs, squared correlation, Chi-square p-values, and t-test p-values for both the discovery and verification groups are provided in (**Supplementary Data file 10**).

### Candidate Bivariate Marker Selection

Feature selection of the 110 verified bivariate ratios was performed using an XGBClassifier model from the python package XGBoost (v. 1.4.0). The test size was set at 0.15, random state equaled 42, and feature importance values were obtained from the coefficients. The bivariate found to have the largest feature importance was used to sort the 110 bivariates and all samples higher than 90% of the control samples along with the bivariate were removed for the second iteration of the XGBClassifier. The process was repeated for a total of three iterations/bivariates. Principal component analysis of all the samples using the three highest bivariates was performed using PCA from sklearn (v. 0.23.2) and plotted using plotly (v. 4.7.0). Sensitivity was calculated as true positives/(true positives + false negative) and positive predictive value was calculated as PPV = (sensitivity × prevalence)/ ((sensitivity × prevalence) + ((1 – specificity) × (1 – prevalence))) where prevalence in the study was equal to 57.8% and specificity was 78.8%.

### Statistical Analysis

Clinical data were analyzed using Students t-test, Mann-Whitney U Test and Fisher’s Exact Test with (SPSS version 25). Hierarchical clustering was performed and displayed using the Qlucore Omics Explorer Software (Qlucore, Lund, Sweden) to visualize candidate miRNAs identified using the PEER process. Clustering and enrichment of the 110 bivariate biomarkers was done using the cumulative distribution function and is detailed above.

## Results

### A. Characteristics of the Study Population

A total of 131 subjects were recruited and enrolled. Of these, 1 subject was lost to follow-up and 7 were excluded during quality control of the small RNA sequencing data due to insufficient miRNA complexity or low read depth (**Figure 1A**). Cases were defined as pregnancies with an adjudicated PE diagnosis (n=71) and controls (n=52) were from participants with either hypertension that did not meet the criteria for PE or normal (nonhypertensive) pregnancy outcomes. The pass-filter 71 cases and 52 controls were subjected to further analysis. Cases and Controls were divided into Discovery and Verification cohorts at a ratio of 2:1. The Discovery and Verification cohorts were matched by fraction of Cases and Controls (**Figure 1B**) and Gestational Age at Blood Draw (GABD). **Supplementary Data file 1** lists all subjects and includes assignment to discovery and verification cohorts, assignment as cases and controls, and detailed adjudicated diagnoses.

**Fig. 1.**
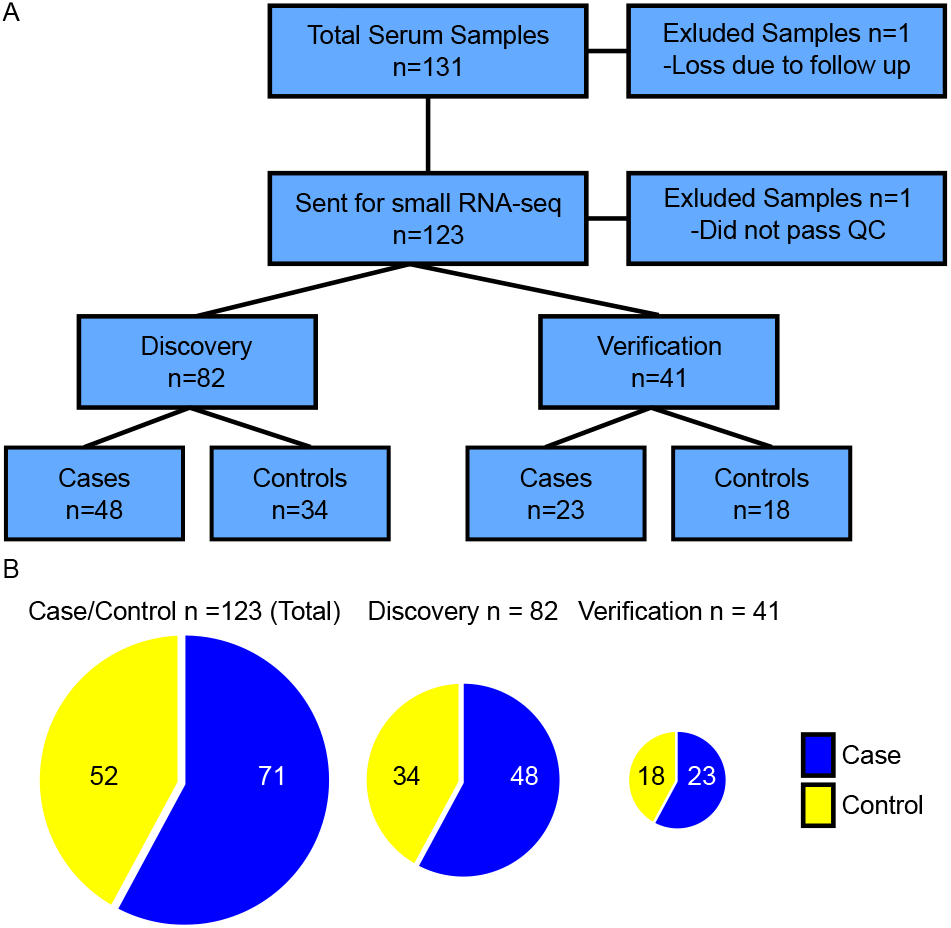
Study Population Characteristics: (A) Flow Chart depicting the exclusion of 7 subjects due to insufficient RNA quality and 1 subject due to loss at follow up. Remaining 123 samples were submitted for analysis (71 cases, 52 controls). (B) Pie charts depicting the breakdown of cases and controls in the cohort.

Comparisons for demographic and clinical factors between the Discovery and Verification cohorts, and between the Cases (PE Dx) and Controls (All Other Dx) were then performed (**Table 1**). Significant differences (p-value *≤* 0.05) were observed for median GA at delivery, mean birth-weight, small for gestational age (SGA, birth weight of < 10%), and admission to the NICU between the Cases and Controls in both the Discovery and Verification cohorts; these differences are expected given the known associations between PE and outcomes such as iatrogenic delivery and SGA neonates. Interestingly, there was also a significant difference in race/ethnicity between Cases and Controls, with higher percentages of Hispanic and non-White subjects in the Case compared to the Control category. All other factors were similar between Cases and Controls for both the Discovery and Verification Cohorts. The only significant difference between the Discovery and Verification Cohorts was seen for Race/Ethnicity, with the predominant difference being a larger proportion of White non-Hispanic participants in the Discovery cohort.

**Table 1:**
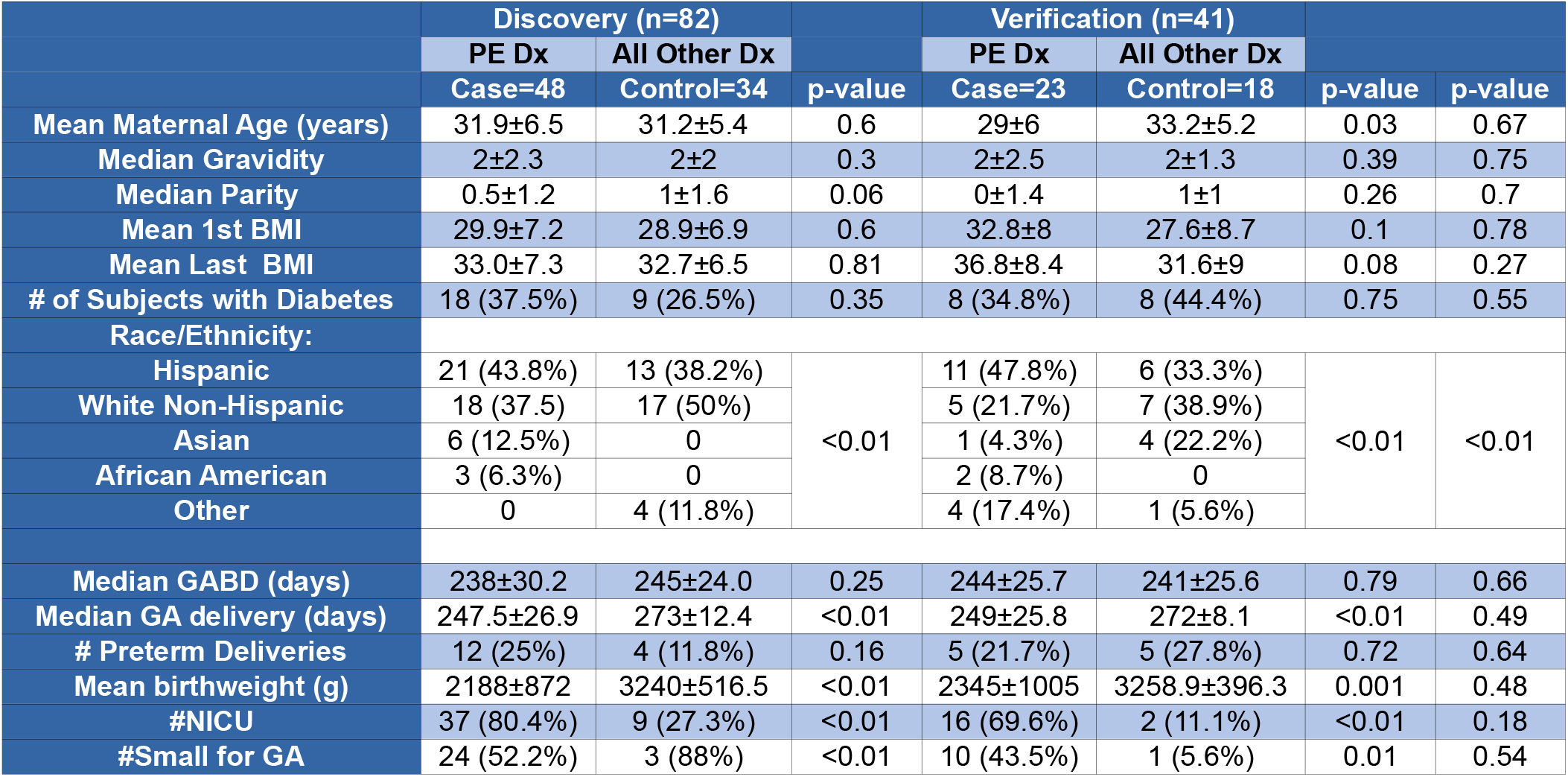
Study subject demographics and clinical characteristics. Demographics and clinical characteristics of subjects used for statistical analysis. p-values represent statistical significance for characteristics between cases and controls within the Discovery and Verification cohorts, and between the Discovery and Verification cohorts (rightmost column). Note: GA=Gestational Age, GABD=Gestational Age at Blood Draw, NICU=Neonatal Intensive Care Unit, Small for GA = Birthweight <10^th^ percentile for GA.

### B. Generation and Pre-Processing of ex-miRNA Data

As detailed in the Methods, exRNA was extracted from maternal serum samples and then subjected to small RNA sequencing. The small RNA-seq data were trimmed, cleaned, and mapped using the exceRpt pipeline(43), and the raw count data were used for quality control, removing samples with total miRNA counts <500,000 or complexity <300 miRNAs with at least 10 raw counts in at least 50% of the samples. Transformation and batch normalization was conducted using the PEER Package as described in the Methods.

### C. Identification of Univariate ex-miRNA Biomarkers for Early Diagnosis of PE

The PEER Package was used to identify candidate univariate biomarkers comprised of single ex-miRNAs.

The ex-miRNAs were ranked according to Chi-square p-value in the Discovery cohort, and the top 100 were then evaluated in the Verification cohort. The top ten univariate ex-miRNAs ranked by Chi-square p-value in the Verification cohort that also had a Discovery Chi-square rank < 100 are shown in **Table 2**. For the Verification cohort, in addition to the p-value, we used PEER to compute the 25^th^ and 75^th^ percentile AUCs for these candidate ex-miRNA biomarkers, which is done by subsampling the samples 1000 times. We discovered that although only a subset of the candidate ex-miRNAs reached statistical significance according to the Chi-square p-value, all of the candidate ex-miRNAs that were more highly expressed in non-PE samples compared to PE samples showed a 75^th^ percentile AUC of <0.35, and all of the candidates that were more highly expressed in PE samples compared to non-PE samples showed a 25^th^ percentile AUC of >0.65.

**Table 2:** Top 10 ranked univariate ex-miRNAs. ex-miRNAs were ranked according to Chi-square p-value in the Discovery cohort, and the top 100 were then evaluated in the Verification cohort. The top ten univariate ex-miRNAs ranked by Chi-square p-value in the Verification cohort that also had a Discovery Chi-square rank < 100 are shown. Discovery and Verification rank and Chi-square p-value are shown along with the Verification AUC at the lower 25^th^ and upper 75^th^ percentiles and whether the expression of the ex-miRNA is upor down-regulated in the non-PE vs the PE samples.

|  |  | <0.05 |  | <0.05 | >0.65 | <0.35 |  |
| --- | --- | --- | --- | --- | --- | --- | --- |
| miRNA | training rank (Chisq pval) | Discovery Chisq pval | Discovery Rank | Discovery Chisq pval | Discovery AUC lower 25 | Discovery AUC upper 75 | Non-PE vs PE |
| hsa.miR.4732.5p | 64 | 0.299 | 4 | 0.039 | 0.008 | 0.2 | Up |
| hsa.miR.363.3p | 22 | 0.188 | 3 | 0.039 | 0.007 | 0.201 | Up |
| hsa.let.7b.5p | 1 | 0.015 | 5 | 0.047 | 0.012 | 0.22 | Up |
| hsa.miR.423.5p | 2 | 0.041 | 2 | 0.036 | 0.011 | 0.221 | Up |
| hsa.miR.144.3p | 75 | 0.317 | 11 | 0.077 | 0.046 | 0.307 | Up |
| hsa.miR.1323 | 82 | 0.326 | 9 | 0.075 | 0.683 | 0.945 | Down |
| hsa.miR.361.5p | 68 | 0.307 | 8 | 0.07 | 0.708 | 0.958 | Down |
| hsa.miR.512.3p | 31 | 0.206 | 10 | 0.076 | 0.725 | 0.961 | Down |
| hsa.miR.518e(519a,519b,519c,522,523).5p | 96 | 0.352 | 7 | 0.063 | 0.745 | 0.975 | Down |
| hsa.miR.516a.5p | 30 | 0.205 | 1 | 0.034 | 0.801 | 0.991 | Down |

The expression of these top ten candidate univariate ex-miRNA biomarkers were then examined on a persample basis (**Figure 2, Supplementary Data file 2**). As can be seen in **Figure 2**, there is a strong separation of the samples by overall diagnosis (Case vs. Control), by detailed diagnoses, and by Interval between blood draw and PE diagnosis, but not by Cohort (Discovery vs. Verification) or GABD. Superimposed PE cases appear to be scattered across several clusters of samples, some of which are comprised mostly of other PE samples and others of hypertensive non-PE samples; this may be due to the fact that it is clinically challenging to diagnose this condition and/or that it contains molecular features of both chronic hypertension and PE. We note that four of these ten candidate miRNAs are encoded on Chr19 and are more highly expressed in PE compared to non-PE, and three are on Chr17 and are more highly expressed in non-PE.

**Fig. 2.**
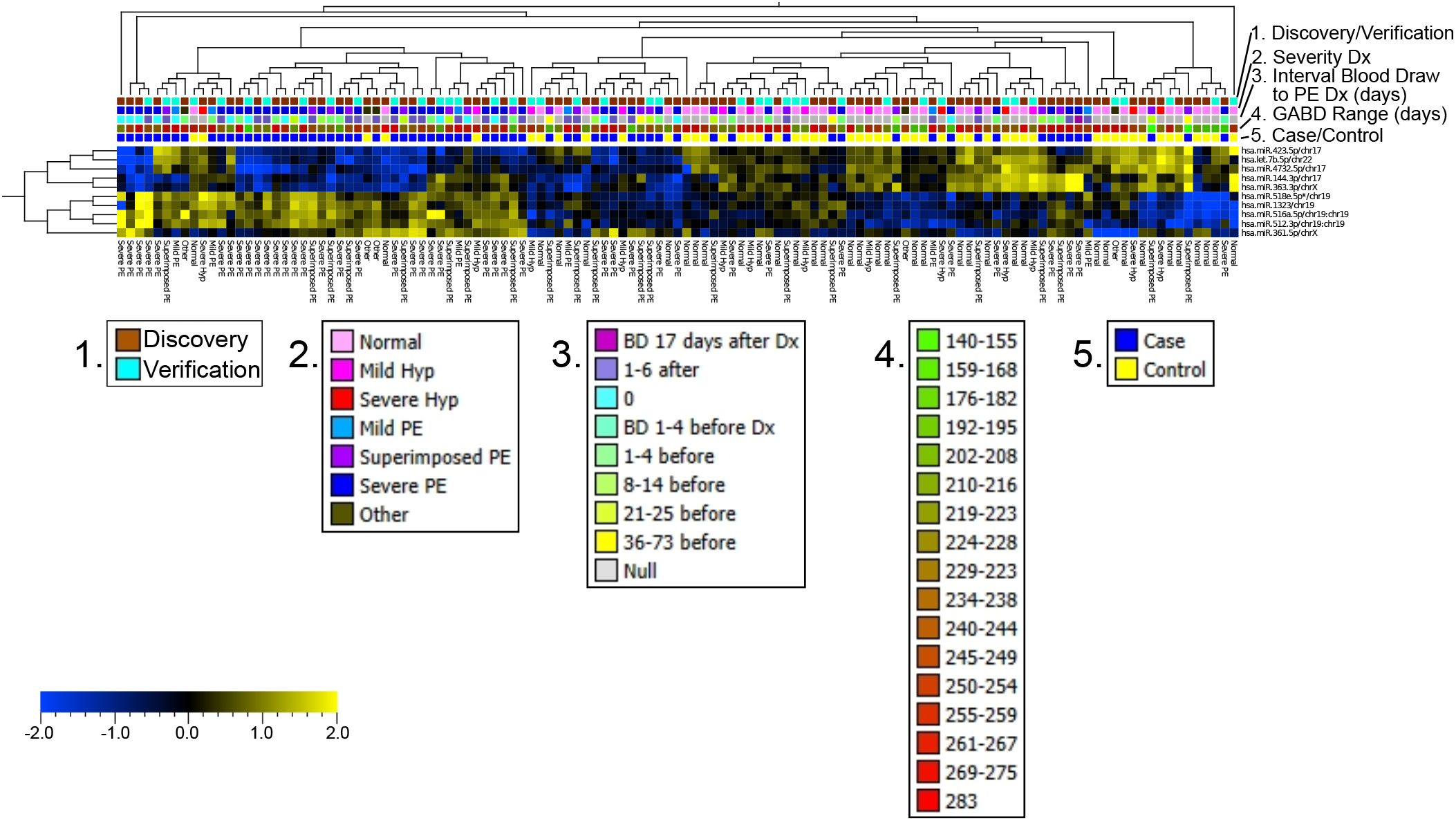
Expression of the top 10 candidate univariate ex-miRNA biomarkers: Heatmap displaying the expression of the top 10 candidate univariate ex-miRNA biomarkers. Both ex-miRNAs (y-axis) and samples (x-axis) are clustered using hierarchical clustering. Color-bars at the top of the heatmap display sample metadata and show strong separation of the samples by overall diagnosis (Case vs. Control), by detailed diagnoses, and by Interval between blood draw and PE diagnosis, but not by Cohort (Discovery vs. Verification) or GABD.

### D. Identification of Bivariate ex-miRNA Biomarkers for Early Diagnosis of PE

The PEER Package was used to identify candidate bivariate biomarkers comprised of ratios of the log values for pairs of ex-miRNAs. To do this, the bivariate features were ranked in the Discovery cohort according to a composite score derived from four metrics: correlation coefficient; Chisquare p-value; 25^th^ percentile AUC; 75^th^ percentile AUC (see Methods for details). The top 1000 bivariate features from the Discovery cohort were then evaluated in the Verification cohort, and the features with a 25^th^ percentile AUC of at least 0.7 were selected, yielding 110 candidate bivariate biomarkers that passed Verification (**Supplementary Data file 3**).

The expression of these top 110 candidate bivariate ex-miRNA biomarkers were then examined on a persample basis (**Figure 3, Supplementary Data file 4**). As can be seen in **Figure 3**, there is strong separation of the samples by overall diagnosis (Case vs. Control), by detailed diagnoses, and Interval between blood draw and PE diagnosis, but not by Cohort (Discovery vs. Verification), GABD, maternal diabetes, or BMI.

**Fig. 3.**
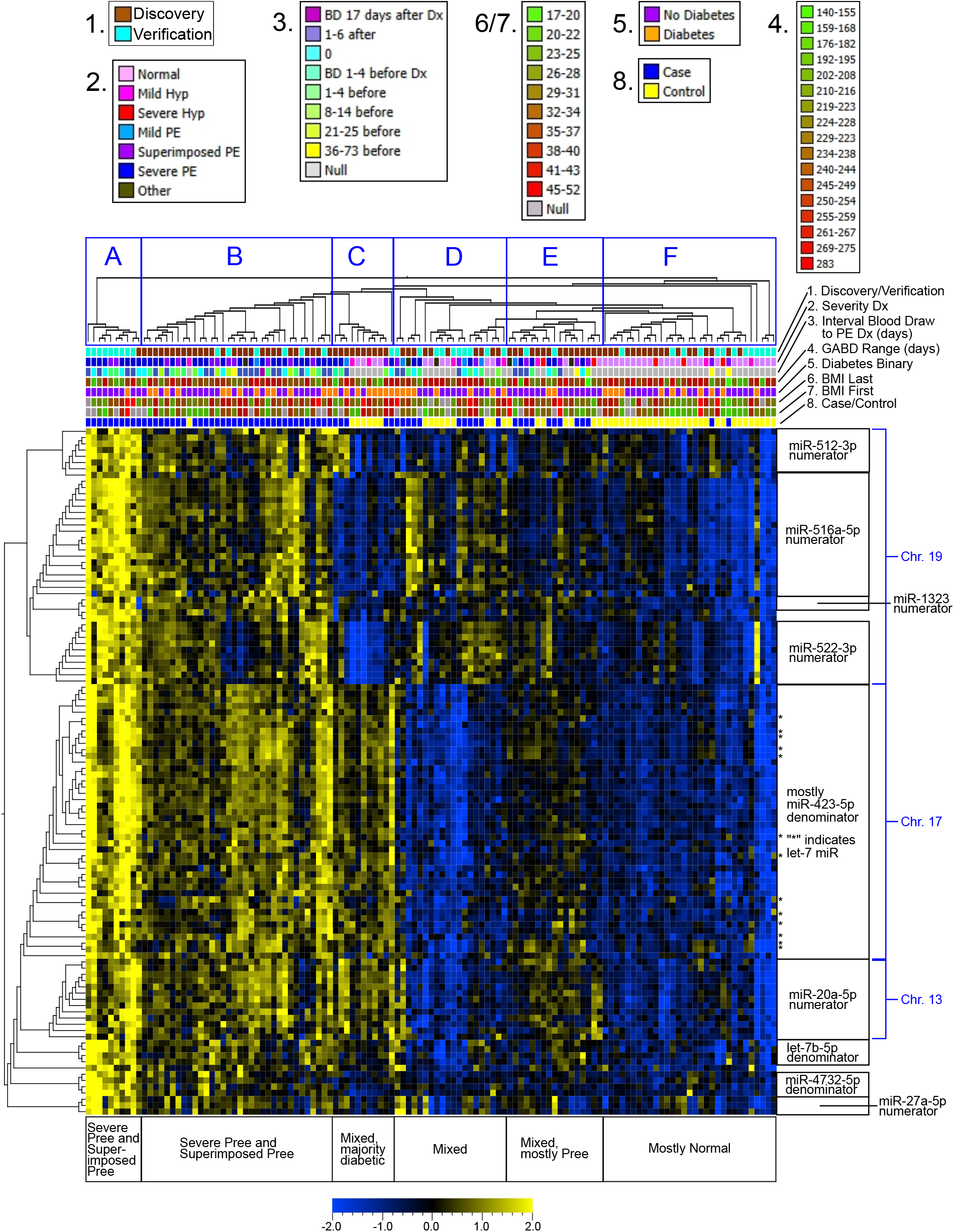
Expression of the 110 verified candidate bivariate ex-miRNA biomarkers: Heatmap displaying the log ratio of the 110 verified candidate bivariate ex-miRNA biomarkers. Both bivariate ex-miRNAs (y-axis) and samples (x-axis) are clustered using hierarchical clustering. Color-bars at the top of the heatmap display sample metadata and show strong separation of the samples by overall diagnosis (Case vs. Control), by detailed diagnoses, and by Interval between blood draw and PE diagnosis, but not by Cohort (Discovery vs. Verification) or GABD. Blue boxes with letters mark sample clusters discussed in the text. Bivariate ex-miRNA biomarkers clustered by the presence of a particular ex-miRNA biomarker in the ratio and by location in the genome as shown on the y-axis.

Inspecting the miRNA composition of these top 110 candidate bivariate biomarkers, we observe that several of the candidate univariate biomarkers are present, and that the Chr19 biomarkers are significantly enriched in the numerators (p-value < 0.01 × 10^28^, CDF of the hypergeometric dist.) and the Chr17 biomarkers are significantly enriched (p-value < 0.01 × 10^36^, CDF of the hypergeometric dist.) in the denominators. Certain individual miRNAs appear in multiple bivariate biomarkers, and these sets of bivariate biomarkers tend to cluster together by hierarchical clustering (**Figure 3**). When visualized by chromosomal location, it is apparent that not only are certain individual miRNAs enriched in the set of 110 candidate bivariate biomarkers, but that certain genomic locations are overrepresented, particularly on chromosomes 17 and 19 (**Supplementary Figure 1**).

Compared to the candidate univariate biomarkers, the candidate bivariate biomarkers appear to better separate subjects into phenotypic subgroups, as indicated by the text in the boxes below the heatmap in **Figure 3**. Going from left to right, **Cluster A** consists mostly of samples from subjects who developed Severe PE, along with 2 cases of Superimposed PE, which have uniformly high expression of all of the candidate bivariate biomarkers. The interval between GABD and initial diagnosis of any form of PE in this group ranges from 14 days before to 6 days after, suggesting that this pattern of biomarker expression indicates a current or imminent clinical diagnosis of PE with severe features. **Cluster B** contains a relatively even mixture of cases of Severe PE and Superimposed PE, along with one case of Severe Hypertension and one case of Mild PE. The samples in **Cluster B** express moderate levels of all the candidate bivariate biomarkers. The interval between GABD and initial diagnosis of any form of PE in **Cluster B** is generally larger than in **Cluster A** and ranges from 73 days before to 6 days after, suggesting that the pattern of biomarker expression in **Cluster B** may serve as an early predictor of PE with severe features. **Cluster C** shows uniform and moderate expression of the bivariate biomarkers in the lower portion of the heatmap (**Figure 3**, miR-20a-5p numerator, miR423-5p denominator, let-7 family) and mostly low expression of the biomarkers in the upper portion of the heatmap (**Figure 3**, miR-512-3p numerator, miR-516a5p numerator, miR-522-3p numerator), and consists of a mixture of Severe, Mild, and Superimposed PE, Mild Hypertension, and Normal cases. However, it does appear that all but one of the PE cases shows moderately high expression of the biomarkers with miR-512-3p in the numerator, suggesting that **Cluster C** is in fact comprised of two subclusters, one for PE, and one for non-PE. **Clusters D and E** contain samples with a broad mix of diagnoses, and a small number of cases of Severe PE, but mostly Mild and Superimposed PE, Mild and Severe Hypertension, and Normal. **Cluster D** samples show low-to-moderate expression of the biomarkers in the upper portion of the heatmap, and essentially no expression of the biomarkers in the lower portion of the heatmap, while **Cluster E** shows low expression of the biomarkers in the lower portion of the heatmap and very low expression of the biomarkers in the upper portion of the heatmap. **Cluster F** shows little to no expression of any of the bivariate biomarkers, and is comprised of mostly Normal samples, with a few Mild and Severe Hypertension and very few Superimposed PE samples. Given that there was a cluster of bivariate biomarkers that was enriched for let-7 family miRNAs in both the numerator and denominator, we examined the expression of the let-7 family miRNAs in our dataset in more detail (**Figure 4A**). We observed that in the top 110 bivariate biomarkers, let-7b-5p, let-7c-5p, and let7e-5p occurred in the denominator, which is consistent with our univariate findings that these miRNAs are expressed at lower levels in PE compared to non-PE samples. Although the 3p/5p arms are not perfectly concordant between the bivariate and univariate datasets, the let-7a/d/f/g/i miRNAs are consistently found in the numerators and more highly expressed in PE than non-PE. When we examine the sequences of these miRNAs, we find that for the 5p sequences, a C in position 18 is associated with higher expression in PE as a univariate biomarker (**Figure 4B**)/bivariate numerator (**Figure 4D**), while a U in position 18 is associated with lower expression in PE as a univariate biomarker (**Figure 4C**)/bivariate denominators (**Figure 4E**). The 3p sequences were only found to have higher expression in PE as univariate biomarkers (**Figure 4B**)/bivariate numerator (**Figure 4D**); we note that the sequence concordance among these let-7-3p family members was markedly lower than for the 5p family members.

**Fig. 4.**
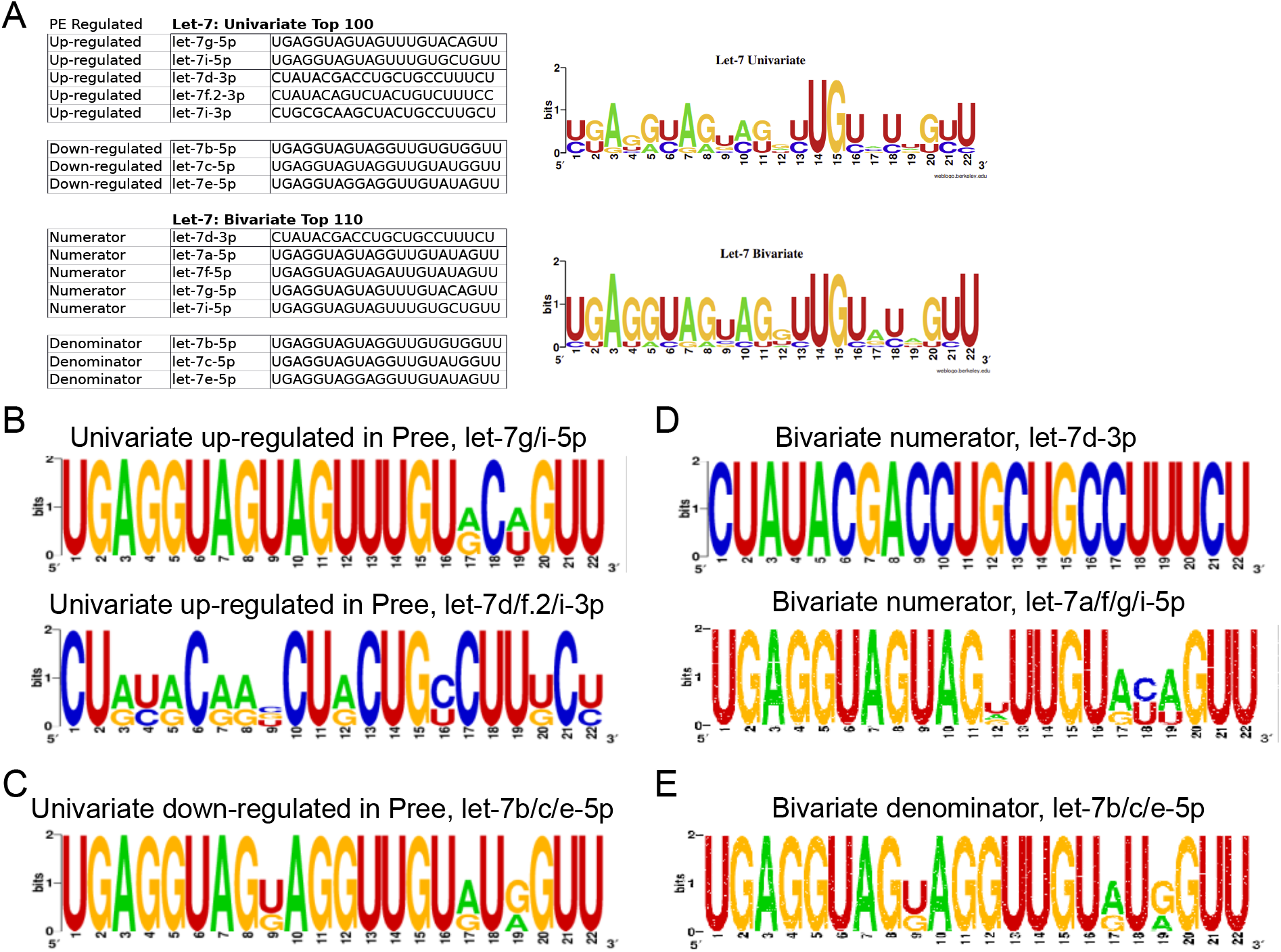
Let-7 family clusters and sequence logos. (A) Table showing the let-7 family ex-miRNAs found in the top 100 univariate biomarkers (top) and the top 110 bivariate biomarkers (bottom). The univariate table is divided by let-7 ex-miRNAs that are either up-regulated or down-regulated in the PE samples and by 5p and 3p. The bivariate table is divided by the let-7 ex-miRNAs found in either the numerator or the denominator of the bivariate ratio. The sequence logos to the right of the table show a graphical representation of the sequence conservation of nucleotides in the ex-miRNAs. (B) Sequence logos showing a graphical representation of the two let-7 univariate 5p sequences (top) and the three let-7 3p sequences (bottom) up-regulated in our preeclampsia samples. (C) Sequence logo showing a graphical representation of the three let-7 univariate 5p sequences down-regulated in our preeclampsia samples. (D) Sequence logos showing a graphical representation of the one let-7-3p (top) and four let-7 5p (bottom) sequences found in the numerator of the bivariate ratios. (E) Sequence logo showing a graphical representation of the three let-7-5p sequences found in the denominator of the bivariate ratios.

### E. Expression patterns of candidate biomarkers across detailed diagnoses

Given that we had detailed adjudicated diagnoses available (Severe PE =PE with severe features, including HELLP), Superimposed PE, Mild PE =PE without severe features), other (Atypical PE, Mild PE with SGA, Severed gestational hypertension with SGA, Gestational proteinuria, Gestational thrombocytopenia, Severe chronic hypertension with headache and SGA), Mild hypertension (Mild chronic hypertension or Mild gestational hypertension), Severe hypertension (Severe chronic hypertension or Severe gestational hypertension), and Normal), we examined the expression of our candidate univariate and bivariate biomarkers across these more granular categories. Ordering the detailed diagnoses from more severe to less severe (i.e. Severe Preeclampsia on the left to Normal on the right), we see, as might be expected that for most of the ten candidate univariate biomarkers that there is a smooth progression from either higher to lower, or lower to higher, expression (**Supplementary Figure 2**). However, interestingly, there were a few miRNAs for which the levels in samples from Severe Hypertension are more similar to Normal and Mild Hypertension are more similar to PE (let-7b-5p, miR-1443p, miR-361-5p, miR-423-5p).

For the 110 candidate bivariate biomarkers, we performed hierarchical, agglomerative clustering, using a weighted method with a Pearson correlation distance metric to identify ten different patterns of expression across the detailed diagnoses. The patterns for three of the largest clusters are shown in **Figure 5A-C**. Interestingly, the majority of miRNAs in these three clusters are found in just two regions on the genome, one on chromosome 17 and the other on chromosome 19. Cluster 2 and cluster 3 (**Figure 5A-B**) were significantly enriched for miRNAs found on chromosome 19 in their numerator (p-value < 0.005 CDF of the hypergeometric dist.). 90% of the miRNA’s found in the numerator of cluster 2 and all of the miRNAs in the numerator of cluster 3 come from the primate-specific microRNA cluster C19MC. The largest cluster, cluster 7 (**Figure 5C**), was significantly enriched (p-value < 0.00005, CDF of the hypergeometric dist.) for the chromosome 17 miRNA miR-423-5p, found in the denominator of over 80% of bivariates in the cluster. This miRNA is in the same cluster as miR-4732-5p, which was enriched in cluster 2 (**Figure 5A**). Compared to both the normal and severe hypertension groups, this cluster contains high ratios of all the PE groups as well as the mild hypertension group.

**Fig. 5.**
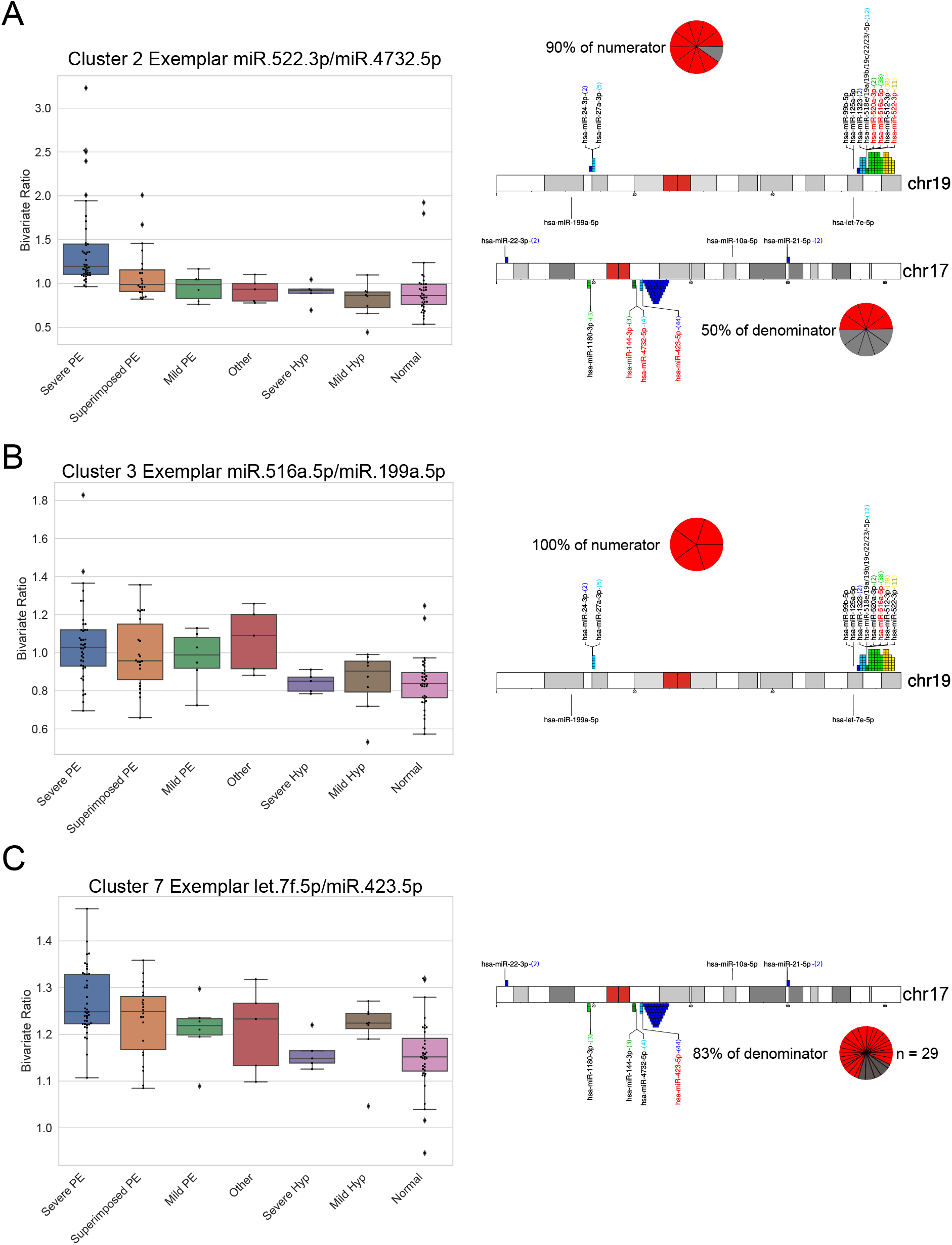
Clustering of the 110 verified candidate ex-miRNA biomarkers. Three of the largest clusters (out of ten) formed using hierarchical, agglomerative clustering using a weighted method and Pearson correlation distance metric. Boxplots (left) show the ratios of an exemplar ex-miRNA. Samples are grouped along the x-axis by their detailed diagnosis. Lines across boxes represent the median, boxes contain the 25^th^ to the 75^th^ percentile. Chromosome ideogram with locations of miRNA clusters found in the 110 verified candidate biomarkers (right). Colored numbers in () at the end of the miRNAs represent the number of ex-miRNAs in the 110 verified ratios potentially present at that location. miRNAs in red font are those present in the cluster displayed in box plot. Pie graphs show the percentage of the clusters numerator or denominator in the displayed chromosome. (A) Boxplot of cluster 2 exemplar miR-522-3p/miR-4732-5p (left) and location of a percentage (pie chart) of cluster 2’s ex-miRNA bivariates (right). (B) Boxplot of cluster 3 exemplar miR-516a-5p/miR-199a-5p (left) and location of a percentage (pie chart) of cluster 3’s ex-miRNA bivariates (right). (C) Boxplot of cluster 7 exemplar let-7f-5p/miR-423-5p (left) and location of a percentage (pie chart) of cluster 7’s ex-miRNA bivariates (right).

Using a recently reported approach for estimation of the fractional contributions of cell/tissue sources to miRNAs in maternal serum(44), we identified the most likely cell/tissue sources for our 10 candidate univariate and 110 candidate bivariate biomarkers. As shown in **Figure 6**, the large majority of these candidates likely originate from the liver, placenta, platelets, or RBCs. Interestingly, most of the miRNAs attributed to the placenta are in the “Up in PE”/Numerator categories, compared to the other three major cell/tissue sources, where the “Up in PE”/Numerator and “Down in PE”/Denominator categories are quite evenly balanced. The miRNAs that were upregulated in PE were found only in the placenta and to a lesser extent the liver, whereas those that were deemed to be upregulated in our control patients were found in platelets or red blood cells. These analyses suggest that specific miRNA clusters influence pregnancy-related functions and that they are dysregulated in PE placentas. Moreover, the dysregulation of miRNAs can be traced to the placenta or to sources in contact with the maternal blood supply.

**Fig. 6.**
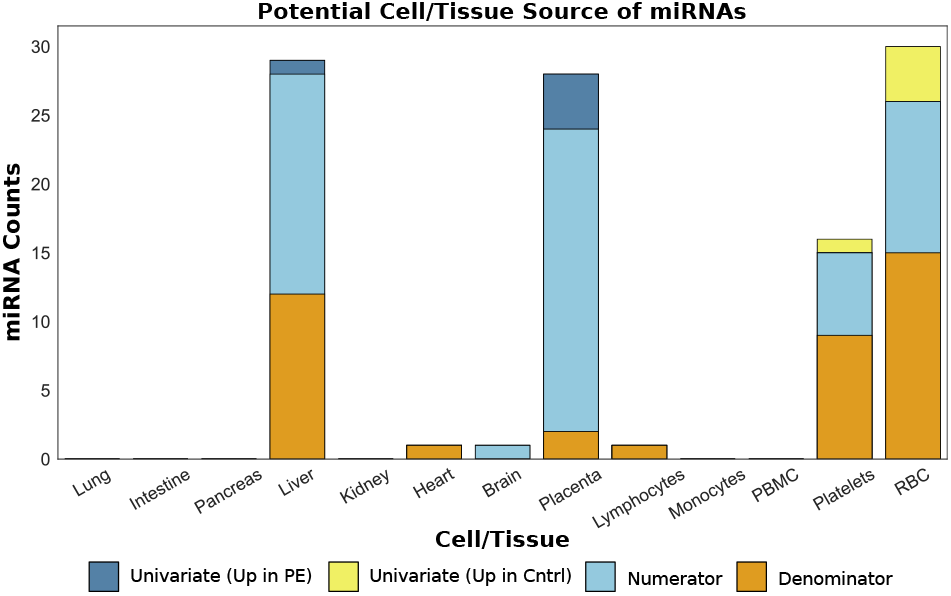
Potential cell/tissue sources for our candidate ex-miRNA biomarkers.

### F. Identification of a small panel of candidate bivariate biomarkers for improved discrimination between cases and controls

To improve discrimination between cases and controls, we identified a small panel of candidate bivariate biomarkers from the set of 110 candidate bivariate biomarkers from the verification cohort using extreme gradient boosting. We performed an iterative approach where we selected the top bivariate, in terms of importance, when running the algorithm on the entire dataset and then removed all of the samples that had a bivariate ratio that was greater than 90% of the controls. We then re-ran the algorithm an additional two times, correctly identifying all but five PE cases, three of which were diagnosed as superimposed PE and one mild PE (**Figure 7A**). The bivariate with the highest importance in our first iteration was miR-522-3p/miR-4732-5p (**Figure 7A**, top heatmap). This bivariate was interestingly also the exemplar bivariate for cluster 2 (**Figure 5A**), which was a cluster of bivariates that clearly separated severe PE patients and normal patients. In fact, this single bivariate was able to separate over 70% of all Severe PE diagnosed cases from controls. The bivariate with the highest importance score on the second iteration was miR-516a-5p/miR-144-3p (**Figure 7A**, middle heatmap). This bivariate contained miR-144-3p, a chromosome 17 miRNA that comprised half of the denominators in cluster 1 (**Figure 5A**) and miR-516a-5p, a chromosome 19 miRNA that comprised 100% of the numerators in cluster 3 (**Figure 5B**), which was a cluster of bivariates that differentiated Superimposed PE cases from controls. The third iteration indicated miR-27b-3p/let-7b-5p had the highest importance score (**Figure 7A**, lower heatmap). This bivariate was found to differentiate Mild PE cases from controls to a larger degree (p-value < 0.0015, t-test) than all but one of the other 110 verified bivariate biomarkers and was responsible for selecting over half of the Mild PE cases from the remaining cases and controls. This iterative panel of three bivariate biomarkers achieved a positive predictive value of 55% at a sensitivity of 93% given a prevalence of pre-eclampsia of 57.7% in our study. We noted that the gestational age and BMI did not appear to be driving the selection of any of three bivariate biomarkers (**Figure 7A, Supplementary Data file 5**). Additionally, we checked these three bivariate markers for correlation with patients’ urine protein-to-creatinine (PC) ratio, 24-hour urine protein levels, platelet count, uric acid, AST, ALT, and systolic and diastolic blood pressure (highest values on the day closest to the date of blood draw) and found very little correlation (maximum correlation < 0.39). We next performed principal component analysis using these three selected bivariate biomarkers (**Figure 7B**) and a randomly chosen set of three bivariate biomarkers from the top ranked 1000 bivariate biomarkers (**Figure 7C**). We found that, as expected, the three selected biomarkers clearly separated the cases and controls on the first three principal components, in the order that they were selected. The first three principal components of the random three biomarkers did not appear to differentiate between cases and controls. To verify that this iterative panel of three bivariate biomarkers would work on a separate cohort we used the ratio cutoff values iteratively for each of the three bivariate biomarkers on a separate cohort of 11 cases and 7 controls processed separately through PEER. This independent validation cohort had a positive predictive value of 85% at a sensitivity of 91% (**Supplementary Figure S3**). These results suggest that a small panel of bivariate biomarkers can be used in an iterative fashion for accurate early detection of patients in the process of developing clinical preeclampsia and can also provide prognostic information related to the severity of the final diagnosis.

**Fig. 7.**
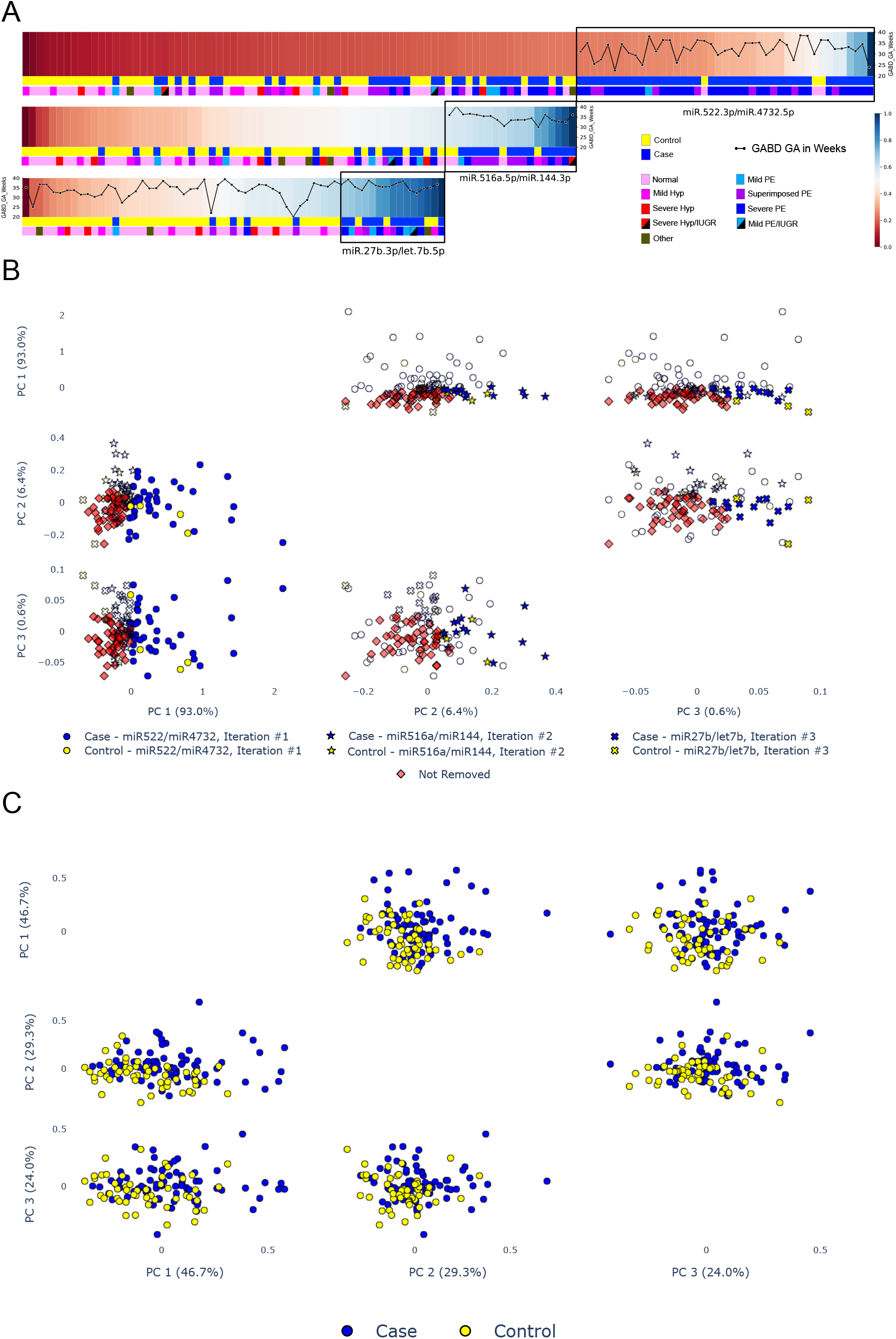
Panel of three candidate bivariate biomarkers to discriminate between cases and controls. (A) Heatmaps showing normalized ratio of each of the three bivariates selected using iterative machine learning approach (see methods). Black boxes indicate samples selected after each iteration (bivariate). Colored bars at the bottom of each heatmap display diagnosis and detailed diagnosis of samples in heatmap. Line graph superimposed on top of heatmap displays the gestational age in weeks at blood draw. (B) Principal component analysis (first three principal components) of the three selected bivariate biomarkers. Blue (cases) and yellow (controls) dots show samples removed in first iteration (left column), blue (cases) and yellow (controls) stars show samples removed in second iteration (middle column), blue (cases) and yellow (controls) x’s show samples removed in third iteration (right column). Red diamonds are samples not removed during iteration and white symbols show cases that were removed during a different iteration. (C) Principal component analysis (first three principal components) of three randomly selected bivariates from the top 1000 ranked bivariates. Blue dots show cases and yellow dots show controls.

## Discussion

Preeclampsia is an important cause of fetal and maternal morbidity and mortality, for which there are currently no highly accurate methods for early diagnosis or prognostic assessment. In this study, we used an unbiased transcriptomic approach to identify and verify univariate (single miRNAs) and bivariate (pairs of miRNAs) extracellular miRNA biomarkers for the early diagnosis of PE in Discovery and Verification cohorts of pregnant women being evaluated in a triage unit for signs and symptoms of PE. Consistent with our previous study focused on prediction of preeclampsia in an *asymptomatic* cohort of pregnant women(44), we were able to verify a larger number of bivariate biomarkers compared to univariate biomarkers, presumably due to the mutual normalizing effect of the miRNA pairs that accounted for the biological and technical variability inherent in small RNA-seq analysis of human biofluid samples. Moreover, we found that in addition to separating cases who developed PE from controls who did not, these ex-miRNA biomarkers could be used to distinguish among different categories of hypertensive disease in pregnancy.

Many prior studies in this area have focused on miRNA known to be enriched in the placenta compared to other organs. However, given that PE is associated with dysfunction of maternal organs, as well as the placenta, we used an unbiased comprehensive transcriptomic approach that enabled us to interrogate maternal serum miRNAs without regard to the source tissue. 123 patients were enrolled from an obstetrical triage unit, where they were being evaluated for signs and/or symptoms of preeclampsia and followed until delivery, to determine whether they were diagnosed with PE prior to delivery (cases) or not (controls). The resulting 71 cases and 52 controls were divided into matched discovery and verification cohorts. Importantly, the cases included subjects with a range of manifestations of PE, and controls included both non-hypertensive subjects and subjects with chronic hypertension and mild gestational hypertension. Cases and controls were well matched in terms of clinical factors, apart from race/ethnicity and differences that were expected given the known associations between PE and outcomes such as iatrogenic delivery and fetal growth restriction, such as median GA at delivery, mean birthweight, small for gestational age, and admission to the NICU.

We identified the top 100 univariate ex-miRNA biomarkers in the Discovery cohort, re-ranked them using the Verification cohort, and selected the top 10 of these for further examination. We found that performing clustering using these 10 univariate biomarkers separated not only cases and controls, but also distinguished between detailed diagnoses and correlated with the interval between blood draw and PE diagnosis. The top 10 included miR-144-3p, miR-1323, miR-518e-5p, and miR-516a-5p, which have been reported in prior ex-miRNA studies to be associated with preeclampsia(25, 30, 31, 34, 38, 40, 41). The two univariate candidate PE biomarkers that were statistically significant in both the discovery and verification cohorts were let-7b-5p and miR-423-5p. Both ex-miRNA biomarkers were found to be at lower levels in our PE cases compared to controls. For let-7b-5p, our results are consistent with Gunel et al., who also reported that let-7b-5p was present at lower levels in the plasma of PE cases compared to controls(27). However, miR-423-5p has been reported in prior studies to be upregulated in maternal plasma in the first trimester of women who later developed early severe PE(36, 47) and in women diagnosed with PE(48). Guo et al. also reported that miR-423-5p was expressed at higher levels in the placentas of patients with PE compared to controls and that it inhibits trophoblast migration, invasion, and proliferation via IGF2BP1(48).

Our bivariate analysis yielded 110 bivariate ex-miRNA biomarkers that passed both Discovery and Verification criteria. The top bivariate ex-miRNA biomarkers by frequency and their appearance in previous reports are shown in **Supplementary Data file 6**. Two of our top ranked miRNA univariate biomarkers, miR-423-5p and miR-20a-5p, were also over-represented in our verified set of bivariate biomarkers, which also contained a disproportionate number of miRNAs from chromosome 19. As noted above, extracellular miR-423-5p has been reported to be more highly expressed in early pregnancy in patients who later develop PE by other groups(36, 47). Our own group also identified this miRNA as the numerator of an early predictive bivariate biomarker for PE(44); we used the same convention in this prior study, where numerators were more highly expressed in PE. This raises the possibility that miR-423-5p may be expressed differently in the maternal circulation in presymptomatic and symptomatic PE.

miR-20a has been previously linked with PE and has been reported to inhibit proliferation, migration, and invasion of JEG3 choriocarcinoma cells by targeting FOXA1(49). As noted above, our verified bivariate biomarkers are enriched for miRNAs encoded in a region on chromosome 19 commonly referred to as the C19MC cluster. Aberrant expression of miRNAs in the C19MC has been previously correlated with PE(28, 50), although the three most highly enriched miRNAs among our verified bivariate biomarkers, miR-512, miR-516, and miR-522, have not been previously linked with PE. The C19MC has been found to be particularly highly expressed in the placenta and in human pluripotent stem cells(51), and has recently been reported to suppress genes critical for maintaining the epithelial cytotrophoblasts stem cell phenotype, and linked to the expression of several genes involved in cell migration and proliferation(52, 53). It has been suggested that dysregulation of this miRNA cluster may result in impaired invasion associated with the shallow placentation of preeclampsia(52).

A markedly higher number of bivariate biomarkers passed in both Discovery and Verification phases of our study compared to univariate biomarkers. The bivariate biomarkers not only showed better separation of cases and controls, but were able to more clearly distinguish among subjects in phenotypic subgroups. We suggest that the superior performance of the bivariate biomarkers may be due to the internal normalization provided by the two miRNAs in each bivariate biomarker to account for technical and biological variability.

In post-hoc analysis, we used a hierarchical, agglomerative clustering package using a weighted method and a custom Pearson correlation distance metric to cluster our Verified bivariate biomarkers into 10 clusters, which not only displayed different patterns of expression across phenotypic subgroups, but also showed enrichment of miRNAs encoded in similar genomic locations, suggesting that miRNAs transcribed from the same region of the genome are coherently regulated by the physiological differences between the phenotypic subgroups. When we analyzed the potential cell or tissue source of these bivariate biomarkers, we found that they largely originated from the placenta, liver, platelets or blood cells. This was not unexpected, given the fact that dysfunction of several of these cells/tissues is associated with preeclampsia (placenta, liver, and platelets), and that they are either highly vascularized or are components of the maternal blood (and therefore are well sampled by serum, the biofluid used for this study). Consistent with a previous study reporting Discovery and Verification of early *predictive* ex-miRNAs for PE in *asymptomatic* gravidas from our group(44), this current study found that bivariate ex-miRNA biomarkers for early diagnosis of PE in symptomatic gravidas miRNAs were largely comprised of one miRNA from the placenta and one from a nonplacental source, suggesting that miRNAs expressed by non-placental cells/tissues may serve to normalize the placental contributions.

To identify the smallest number of bivariate ex-miRNA biomarkers that could efficiently separate cases from controls, we applied a machine learning approach in an iterative fashion. This consisted of ranking the 110 bivariate biomarkers that passed Discovery and Verification using XGBoost and using to the top-ranked biomarker to identify high-probability PE cases, which were then set aside. XGBoost was then reapplied to the remaining bivariate biomarkers and the remaining cases and controls, two more times. Overall, this iterative process was able to identify over 90% of cases with a false positive rate of less than 10% at each iteration and achieved a positive predictive value of 55% at a sensitivity of 93% given a prevalence of pre-eclampsia of 57.7% in our study. In the first round, the identified cases were predominantly those that developed PE with severe features, as well as a smaller number of superimposed PE cases. The second round identified the majority of the remaining superimposed PE cases and our one severe hypertension/SGA case. The third round identified all but one remaining PE case, which was a mild PE case. Interestingly, the controls misidentified as PE using this process were all patients diagnosed with mild hypertension or gestational proteinuria. No control patients with severe hypertension were misclassified as PE. These results suggest that some patients with mild hypertension or gestational proteinuria may share some physiological features with PE that lead to similar alterations in ex-miRNA expression, whereas severe hypertension may actually be physiologically more distinct from PE. Using a separate cohort of 18 patients, we found all but one PE case and achieved a positive predictive value of 85% at a sensitivity of 91%.

Our approach revealed both univariate and bivariate biomarkers, mapped out a method to predict the development of preeclampsia using three bivariate biomarkers in an iterative manner, and validated this approach in a separate patient cohort. The candidate biomarkers we have found in this cohort of patients will now need to be confirmed on a larger multicenter independent cohort. Future validation studies are required to establish the clinical utility of this approach for early diagnosis and prognosis of PE in the obstetrical triage setting. We believe that validation of some of our candidate biomarkers will allow for better clinical resource allocation, prevent low risk patients from unnecessary admission and procedures, and help develop new therapies for patients at high risk for the development of preeclampsia.

## Data Availability

All data, code, and materials used in the analysis is available through the corresponding author. Data is available in dbGaP under the name Extracellular microRNA Biomarkers for Diagnostic and Prognostic Assessment of Preeclampsia at Triage under the accession number: phs003169.v1.

## List of Supplementary Materials

Data file S1. Case (All Adjudicated PE Diagnosis) /Control (All other subjects, including Hypertensive and Normal Pregnancies).

Data file S2. Normalized expression values of top 10 candidate univariate ex-miRNA biomarkers in all samples.

Data file S3. Top 110 candidate bivariate biomarkers that passed verification.

Data file S4. Normalized expression values of top 110 candidate bivariate ex-miRNA biomarkers in all samples.

Data file S5. Refers to Figure 7.

Data file S6. Overlap between highly enriched bivariate miRNAs in this study and previously reported candidate miRNA biomarkers for PE.

Data file S7. miRNA raw counts and metadata.

Data file S8. Top 1000 bivariate miRNAs with final ranking.

Data file S9. Combined discovery and verification miRNA ratio ranking values for the top 1000 miRNA ratios in the discovery dataset.

Data file S10. Combined discovery and verification univariate miRNA ranking values for all 619 miRNAs.

## Acknowledgements

We thank Marni Jacobs, PhD/MPH and Carolina Luevano-Diaz for their help with identification of cases and controls for the validation cohort, and abstraction of metadata, from the Center for Perinatal Discovery’s Perinatal Biorepository and Database. This study was supported by NIH UH3TR000906. SS was supported in part by NIH T32HD007203. RM was supported in part by NIH T32GM008806. MiSeq and HiSeq Next-Gen sequencing was conducted at the IGM Genomics Center, University of California, San Diego, La Jolla, CA, which is supported in part by NIH P30CA023100. Data management, storage, and analysis were performed using the Extreme Science and Engineering Discovery Environment (XSEDE) Comet at the San Diego Supercomputing Center through allocation MCB140074, as well as the Genboree exceRpt Small RNA-seq pipeline.

## Author contributions

Conceptualization: RM, LP, LCL Methodology: RM, LP, LCL Investigation: RM, LP, SS, CMK, AA, KZR, CT, AH, MM, KV, AM, VT, MM, LLS, MH, GAR, PD, MMP, PP, LCL Visualization: RM, LP Funding acquisition: LCL Supervision: PD, MMP, PP, LCL Writing – original draft: RM, LP, LCL Writing – review editing: RM, LP, SS, CMK, AA, KZR, CT, AH, MM, KV, AM, VT, MM, LLS, MH, GAR, PD, MMP, PP, LCL

## Data and materials availability

All data, code, and materials used in the analysis is available through the corresponding author. Data is available in dbGaP under the name “Extracellular microRNA Biomarkers for Diagnostic and Prognostic Assessment of Preeclampsia at Triage” under the accession number: phs003169.v1.

## Supplementary Figures

**Fig. S1.**
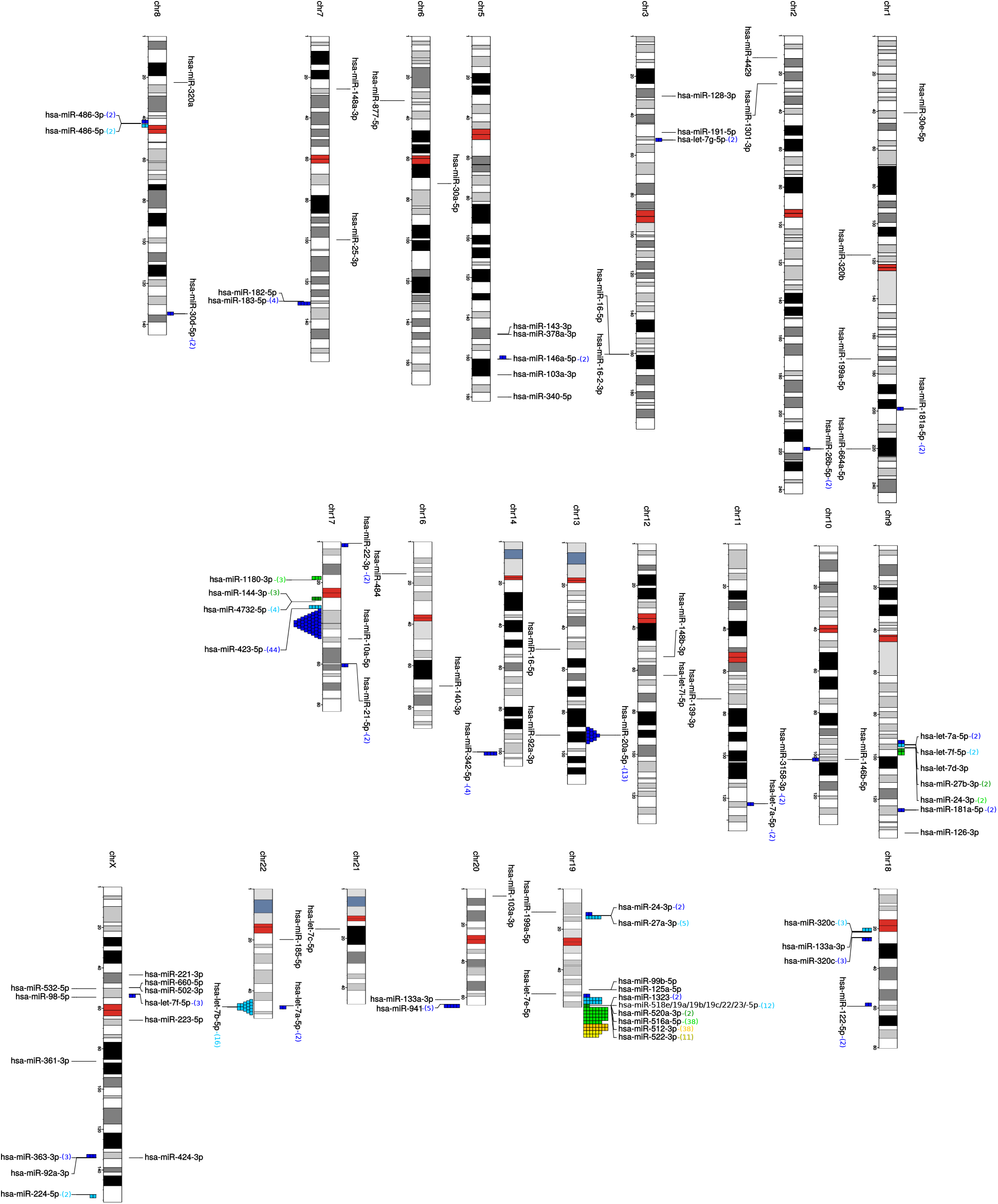
Chromosome ideogram showing the potential of all 110 verified ex-miRNA bivariate biomarkers. Colored numbers in () at the end of the miRNAs represent the number of ex-miRNAs in the 110 verified ratios potentially present at that location. Colored squares on chromosomes give graphical representation of the location of the bivariate candidates.

**Fig. S2.**
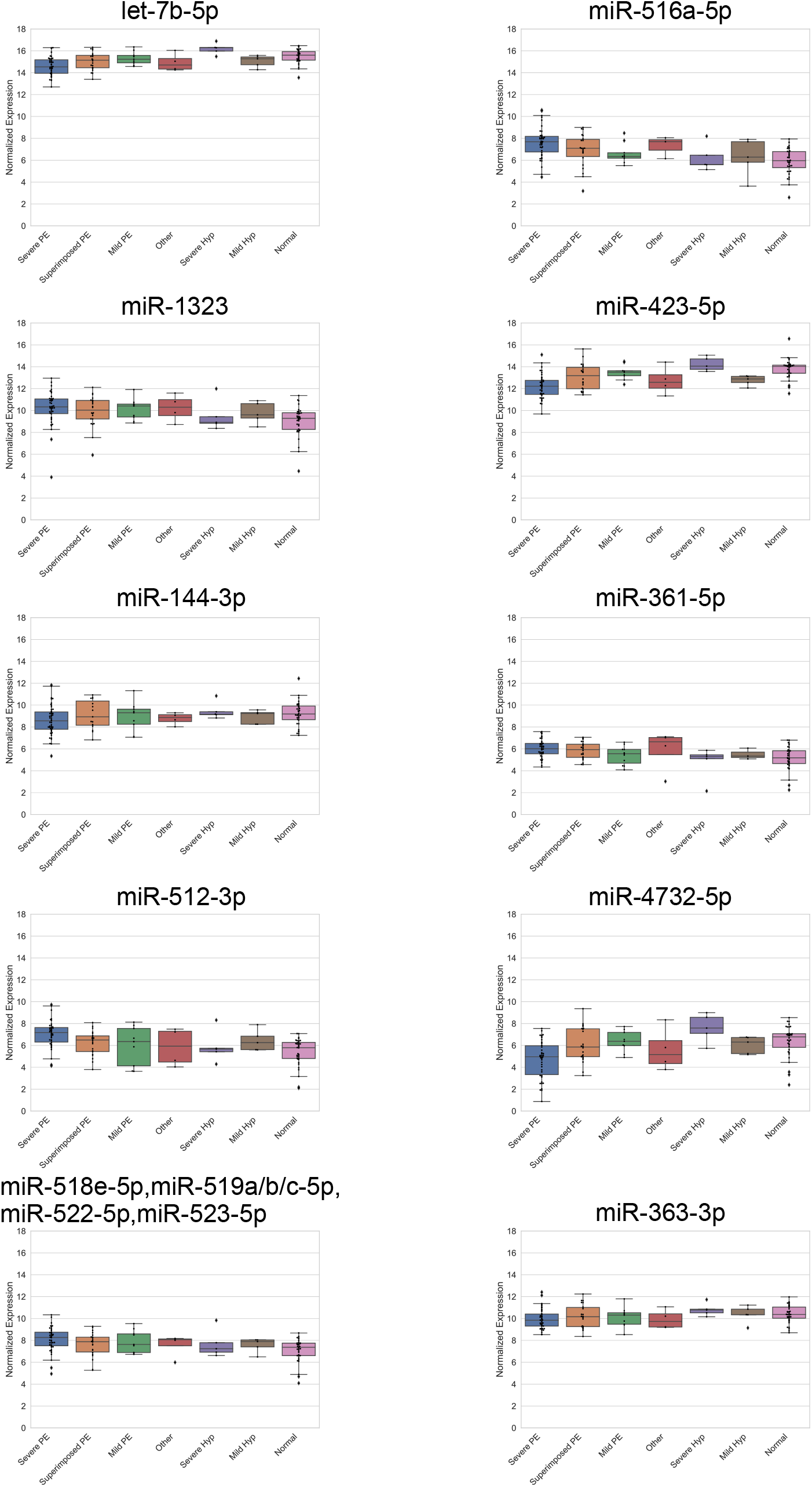
Expression of our ten candidate univariate biomarkers ordered by detailed diagnoses from more severe (left) to less severe (right). Boxplots show the normalized expression of each of the ten candidate univariate biomarkers. Samples are grouped along the x-axis by their detailed diagnosis. Lines across the boxes represent the median, boxes contain the 25^th^ to the 75^th^ percentile.

**Fig. S3.**
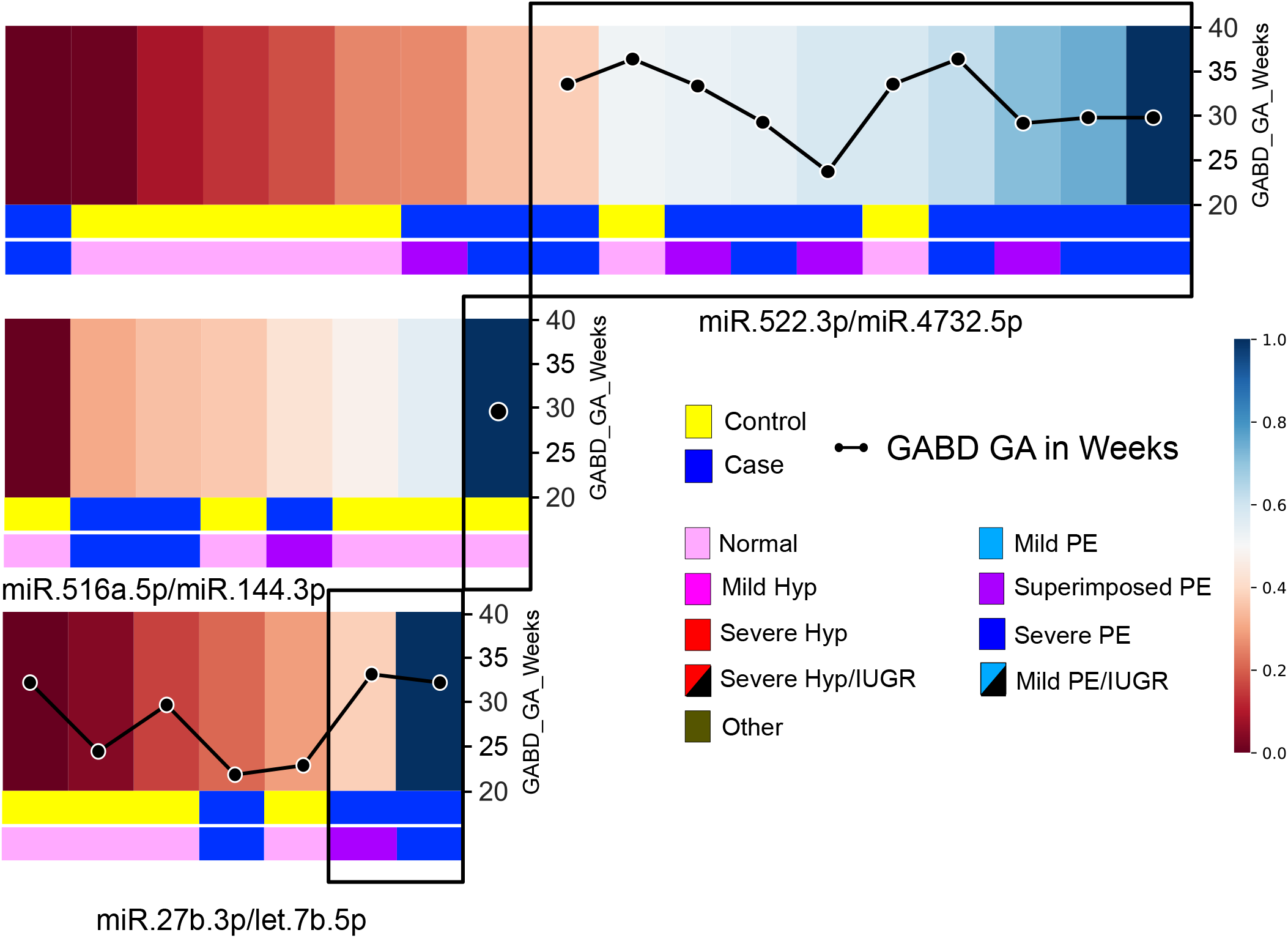
Panel of three candidate bivariate biomarkers to discriminate between cases and controls in independent cohort of 18 patients. Heatmaps showing normalized ratio of each of the three bivariates selected using iterative machine learning approach shown in Figure 7 (see methods). Black boxes indicate samples selected after each iteration (bivariate). Colored bars at the bottom of each heatmap display diagnosis and detailed diagnosis of samples in heatmap. Line graph superimposed on top of heatmap displays the gestational age in weeks at blood draw.

